# TREATMENT OUTCOME AND ITS PREDICTORS AMONG DIABETIC PATIENTS ATTENDING AT SELECTED HOSPITALS OF SOUTHERN ETHIOPIA

**DOI:** 10.1101/2023.07.19.23292871

**Authors:** Bereket Beyene Gebre, Dawit Hoyiso, Kinfe Woldu

## Abstract

**Background:** Diabetes is one of the most serious health crises of the twenty-first century, with the World Health Organization (WHO) estimating that hyperglycemia is the third leading cause of early death globally, following high blood pressure and cigarette use. It is an important cause of blindness, kidney failure, lower limb amputation and other long-term consequences that impact significantly on quality of life. People get sick, die, and health care costs go up as a result.

**Objectives:** Diabetes mellitus treatment outcome and determinants in patients undergoing diabetes treatments in selected hospitals in southern Ethiopia, 2021.

**Methods:** A source population included all diabetic patients who attended chosen hospitals in southern Ethiopia. A cross-sectional study with an institutional focus was conducted in a few hospitals in southern Ethiopia. A pre-tested questionnaire was used to collect information about the patient. Following data cleaning and error checking, the data was loaded into Epi-data to eliminate errors before being exported to SPSS for analysis. The descriptive data were presented using tables and graphics. The result variable was evaluated using a logistic regression model to uncover predictors once the underlying assumptions of the regression analysis were verified. All independent variables with p0.25 were considered for multivariate analysis. The level of significance was then set at p0.05, and the final model was AOR with 95% CL.

**Result:** 277 (65.6%) of the whole sample had positive treatment outcomes. The existence of complication AOR 95%CI; 0.425 (0.217,.832), and elevated fasting blood glucose AOR 95%CI; 0.080 (0.034, 0.188) were revealed to be independent predictors. Given the low size of treatment outcome, providing health information regarding sticking to prescribed medicine and monitoring fasting blood glucose level will result in a positive clinical outcome.

## Introduction

Diabetes mellitus is not a single disease entity, but rather a spectrum of metabolic illnesses characterized by hyperglycemia caused by abnormalities in insulin secretion, insulin action, or both (1). According to the IDF’s 2015 survey, 8.8% of persons aged 20 to 79, or approximately 415 million people worldwide, have diabetes. 75% of these persons lived in low- and middle-income countries. If current trends continue, by 2040, 642 million people (or one out of every ten people) will have diabetes. The most significant gains will occur in regions where economies are transitioning from low-income to middle-income status. In 2015, the IDF projected that 14.2 million persons aged 20 to 79 years had diabetes in Africa, reflecting a 3.2% prevalence (2). Poor diabetes outcome led to complications in many parts of the body and can increase the overall risk of dying prematurely. Heart attack, stroke, kidney failure, leg amputation, visual loss, and nerve damage are all possible consequences. Poorly controlled diabetes raises the risk of fetal mortality and other problems during pregnancy (3). According to the World Health Organization (WHO), hyperglycemia is the third leading cause of premature death worldwide, following excessive blood pressure and cigarette use (4). It causes sickness, death, and an increase in health-care costs. (5). It is the leading cause of end-stage renal disease (ESRD), traumatic lower extremity amputations, and adult blindness. It also predisposes to cardiovascular diseases. With an increasing global prevalence, diabetes will become a prominent cause of illness and mortality in the future. The goal of diabetes treatment is to reduce mortality and complications by stabilizing blood glucose levels. However, despite adequate medication, blood glucose levels may rise, leading in problems such as changes in fat metabolism, nerve damage, and eye disease (4, 6). Diabetes is a major cause of blindness, kidney failure, heart attacks, stroke and lower limb amputation (7).

Ethiopia is among the top four countries with the highest adult diabetic populations in sub-Saharan Africa. The total country prevalence of diabetes mellitus was 5.2%, the number of people with diabetes was 2,567,900, and the number of people with undiagnosed diabetes was 1,960300, according to the IDF 2017 Country Report. Diabetes claimed the lives of 31,000 people, accounting for 81.8% of diabetes deaths among people under the age of 60. Diabetes-related patient attendance and medical admissions in major hospitals have been increasing. In Addis Ababa Ethiopia diabetes-related admissions in have increased from 7% in 2005 to 34% in 2009 (8). This necessitates a shift in healthcare goals as well as current data on treatment outcomes and predictors in Ethiopia. Diabetes complications caused by inadequate sugar management in Ethiopia range from immediate complications such as DKA to chronic complications and mortality.However, there no research evidences in the study area showing treatment outcomes and predictors. Assessing the treatment outcome as good and poor based on glycemic control will provide clinical evidence to bring good health outcome for the patient. So, the aim of this study is to assess treatment outcome and its predictors in the study area in which it provides input for health care providers and institution.

## Objectives

### General objective

To assess treatment outcome and its predictors among DM patients attending treatments at selected hospitals of Southern Ethiopia, 2021

### Specific Objective

To determine treatment outcome of DM patients attending at selected hospitals of southern Ethiopia, 2021

To identify predictors of treatment outcome among DM patients attending treatments at selected hospitals of southern Ethiopia, 2021

## Methods and materials

### Study area and period

#### Study area

The research was carried out at three hospitals: Hawassa University Comprehensive Specialized Hospital, Wolaita Sodo University Teaching Referral Hospital, and Nigist Ellen General Hospital. Hawassa University Comprehensive Specialized Hospital is a teaching hospital located 275 kilometers south of Addis Abeba in Hawassa town, Sidama region. Nigist Ellen Mohammed Memorial Hospital is a general hospital located in Hadiya Zone, 230 kilometers from Addis Abeba. Wolaita Sodo University Teaching Referral Hospital is also located in Sodo town, which is 328 kilometers from Addis Abeba.

#### Study period

The study was conducted from October 24, 2021-February, 30, 2021

### Study design

A cross-sectional study design based on institutions was carried out at chosen hospitals in southern Ethiopia.

#### Source population

All diabetes patients that visited designated hospitals in southern Ethiopia during the study period.

#### Study population

All sampled diabetes patients who attended outpatient department at selected hospitals of southern Ethiopia during the study period.

### Inclusion and exclusion criteria

All diabetic patients who attended outpatient department at selected hospitals in southern Ethiopia who can give informed consent will be included. Whereas, those who are not volunteer have been excluded

### Sample size

Sample size calculation is based on the single population proportion formula inorder to take maximum sample size with no study had done previously involving 3 different regional hospitals in SNNPR, Ethiopia

> **N= [**(Zα /2)^2^ P (1 – P)]/d^2^ is used.

> N**=** is sample size,

> Zα/2 is Z value which is 1.96 at 95% confidence interval,

P is the proportion of diabetes patients, which is 0.5, and d is the marginal error, which is 0.05. In order to accommodate the broadest range of patients visiting diabetic clinics, the sample size will be 384 at 95% confidence interval and p value of 0.5. Adding 10% non-respondent rate, the final sample size will be 422.

### Sampling Technique

From all the hospitals in the southern region of Ethiopia, the research area was chosen at random using a lottery approach. Proportionate sample allocation was done to allocate samples for each hospital of Adare general hospital, Nigist Ellen general hospital and Wolayta sodo general hospital. As a result, every patient who had an appointment during the study period was chosen since patients attending health institutions are chosen at random until the maximum sample size was reached. So, exit interview was conducted at each hospital.

## Variables

### Dependent Variable

Treatment outcome

### Independent Variables

**Socio demographic variables**: - Age, Sex, educational status, marital status, income and job status.

**Behavioral Variables** are: - Alcohol drinking, cigarette smoking, Khat chewing, Physical activity practice, practices to prescribed diet regimen.

**Disease and medication related Variables** are: - Duration of Diabetes, Type of treatment, co-morbid illness, Fasting blood glucose level, DM control, Medication adherence

### Operational definition

**Glycemic control**: patients were categorized based on the American Diabetic Association (ADA) 2017 guideline recommendation [8] into two groups:

**Good treatment outcome**: average fasting blood glucose of 80–130 mg/dL.

**Poor treatment outcome:** average fasting blood glucose of > 130 mg/dL.

### Data collection Instrument

Data abstraction sheets, structured questionnaires, and observation checklists were utilized to collect data from the patient. Investigators created data abstraction sheets after evaluating various literature to extract important information from patient charts and laboratory results. A structured questionnaire was employed for the face-to-face interview. The World Health Organization guideline (WHO), 2012) was used to determine patient. The questionnaires are divided into six sections, which include socio-demographic information, questions about the patient’s physical activity, and questions about the world to determine drug adherence. Laboratory and medication-related data are also included in the questionnaire. The questionnaire also includes laboratory and medication-related information. The information was gathered with the help of 11 BSc Nurses and three M.Sc. health professionals. At the end, the therapy outcome will be measured, which could be excellent or bad.

### Data processing and analysis

Following a physical inspection of the collected data for completeness, the responses were cleaned, edited, coded, and entered into the computer with Epi-data version 3.1. Then the information had been exported to SPSS version 20.0. Before analyzing the data, it had been checked for missing values. Descriptive analysis with frequency had used to investigate the frequency of variables with independent variables. To find predictors of treatment outcome, binary logistic regression with 95% confidence intervals was performed to assess the connection of DM treatment outcome with independent covariates. Finally, a forward stepwise logistic regression model with all independent variables and a p value of 0.25 will be fitted to uncover independent predictors of DM treatment outcome.

### Data Quality

To assure data quality, a well-constructed data collection tool was created, pretested in a hospital that was not chosen to evaluate the instrument’s understandability and applicability, and data collectors and supervisors will receive three days of training. On each data collection day, the supervisor and lead investigator examined and confirmed the collected data for completeness. After double-checking the information, it was entered into Epi-data version 3.1 for double data entry verification.

### Ethical Considerations

In order to follow the ethical and legal standards of scientific investigation, the study was conducted after approval of the proposal by Ethical Review Committee of Hawassa University. Authorities at chosen hospitals granted permission for the study to be carried out. Written informed consent was obtained from each study participant by assuring privacy and confidentiality throughout the data collection period in the Hospital. Individuals who were unwilling to participate from the beginning or at any part of the interview were allowed to withdraw. There was no risk or hazardous procedures putting the participants at harm.

## RESULT

### Socio demographic characteristic of the respondents

A total of 422 diabetes individuals were sampled, obtaining a 100% response rate. There were 276 (62.3%) men and 304 (70.1%) married people among them. The participants’ mean age was 46.51, with a standard deviation of 13.85. (See table 1 below)

**Table 1.**
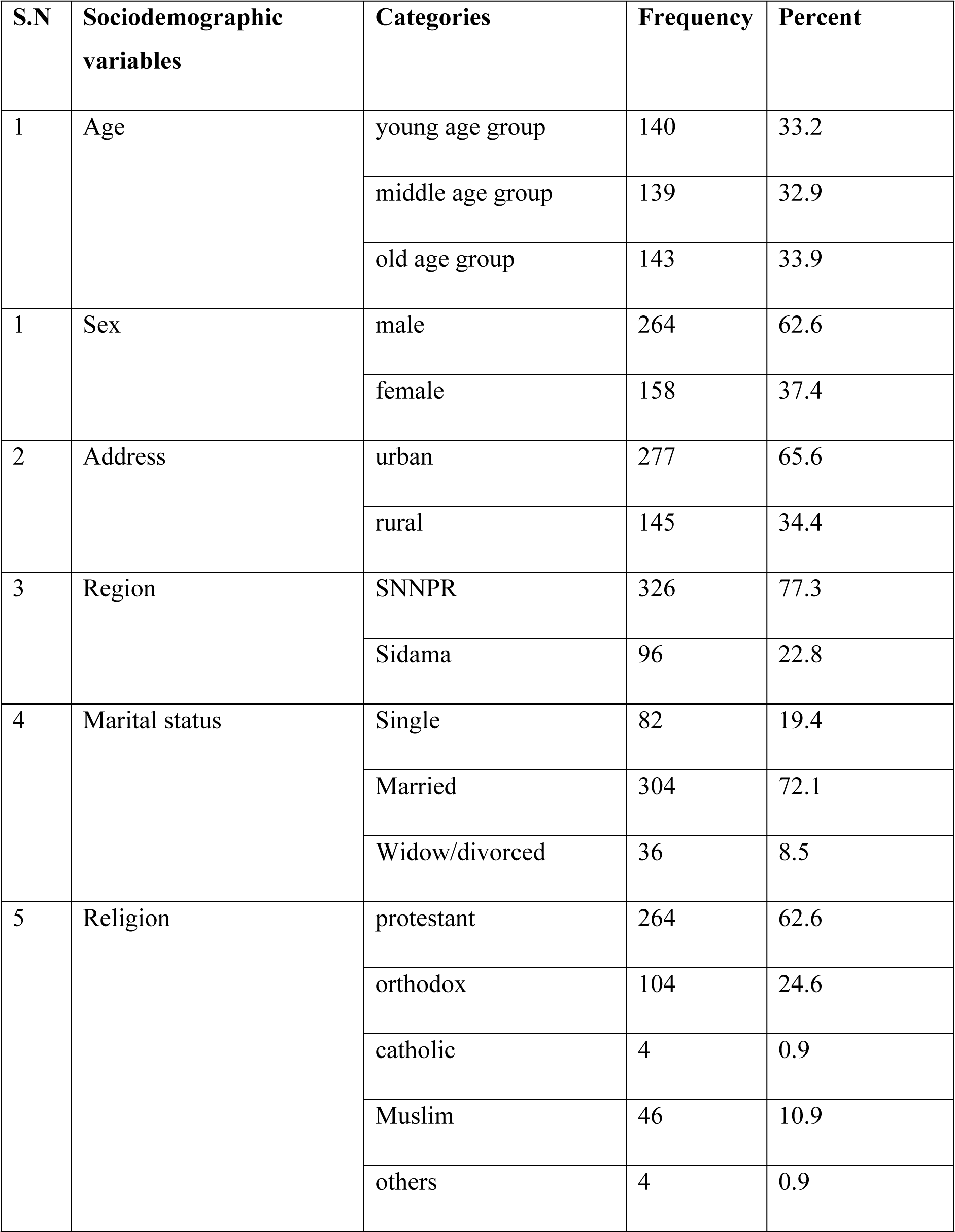
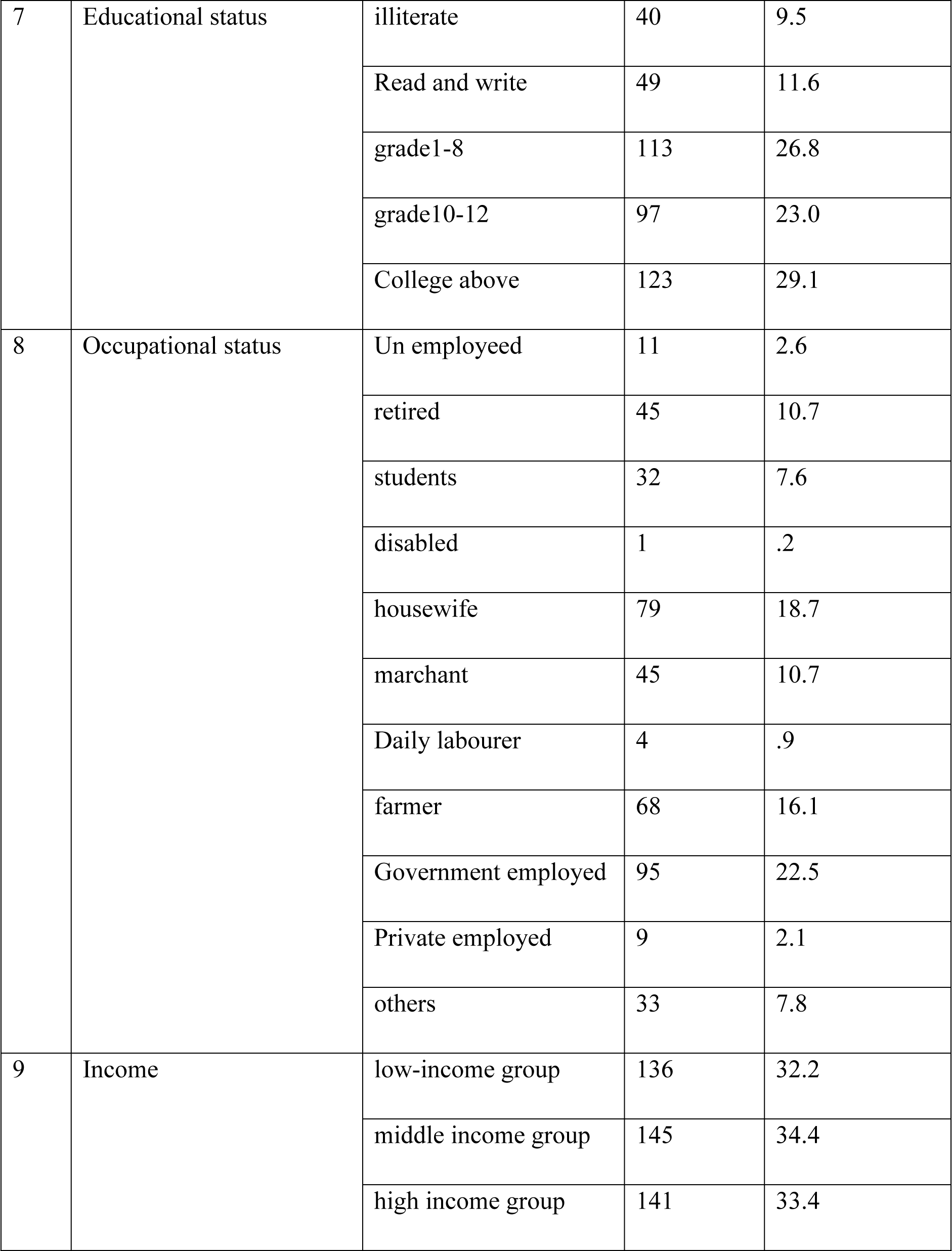
Socio demographic characteristics and factors associated with medication adherence for the respondents at selected hospitals of southern Ethiopia, 2021 (n=422)

| S.N | Sociodemographic variables | Categories | Frequency | Percent |
| --- | --- | --- | --- | --- |
| 1 | Age | young age group | 140 | 33.2 |
|  |  | middle age group | 139 | 32.9 |
|  |  | old age group | 143 | 33.9 |
| 1 | Sex | male | 264 | 62.6 |
|  |  | female | 158 | 37.4 |
| 2 | Address | urban | 277 | 65.6 |
|  |  | rural | 145 | 34.4 |
| 3 | Region | SNNPR | 326 | 77.3 |
|  |  | Sidama | 96 | 22.8 |
| 4 | Marital status | Single | 82 | 19.4 |
|  |  | Married | 304 | 72.1 |
|  |  | Widow/divorced | 36 | 8.5 |
| 5 | Religion | protestant | 264 | 62.6 |
|  |  | orthodox | 104 | 24.6 |
|  |  | catholic | 4 | 0.9 |
|  |  | Muslim | 46 | 10.9 |
|  |  | others | 4 | 0.9 |
| 7 | Educational status | illiterate | 40 | 9.5 |
|  |  | Read and write | 49 | 11.6 |
|  |  | grade1-8 | 113 | 26.8 |
|  |  | grade10-12 | 97 | 23.0 |
|  |  | College above | 123 | 29.1 |
| 8 | Occupational status | Un employeed | 11 | 2.6 |
|  |  | retired | 45 | 10.7 |
|  |  | students | 32 | 7.6 |
|  |  | disabled | 1 | .2 |
|  |  | housewife | 79 | 18.7 |
|  |  | marchant | 45 | 10.7 |
|  |  | Daily labourer | 4 | .9 |
|  |  | farmer | 68 | 16.1 |
|  |  | Government employed | 95 | 22.5 |
|  |  | Private employed | 9 | 2.1 |
|  |  | others | 33 | 7.8 |
| 9 | Income | low-income group | 136 | 32.2 |
|  |  | middle income group | 145 | 34.4 |
|  |  | high income group | 141 | 33.4 |

### Behavioural characteristics and factors

Participants who did not drink alcohol or use cigarettes made up 95.7% and 98.1% of the respondents, respectively. (See table 2 below.)

**Table 2.** Behavioural related characteristics for the respondents at selected hospitals of southern Ethiopia, 2021 (n=422).

### Disease and medication related characteristics

In this study 365 (82.4%) were type II diabetes and 277 (62.5%) had developed complication. Regard to medication adherence and treatment outcome; 195 (44%) and 290 (65.5%) were found to be poor and good adherence and outcome respectively. (See table 3 below)

**Table 3.**
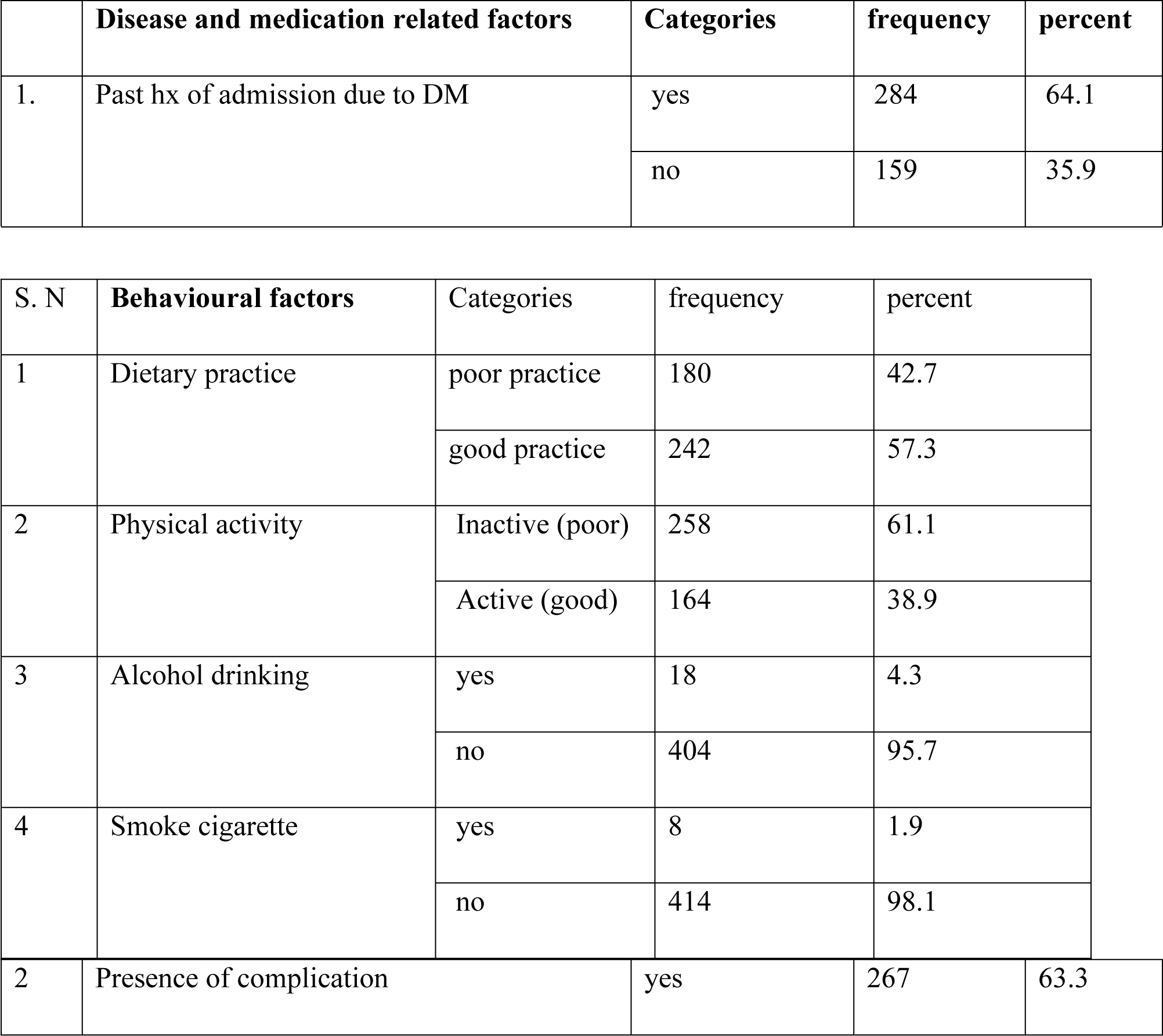
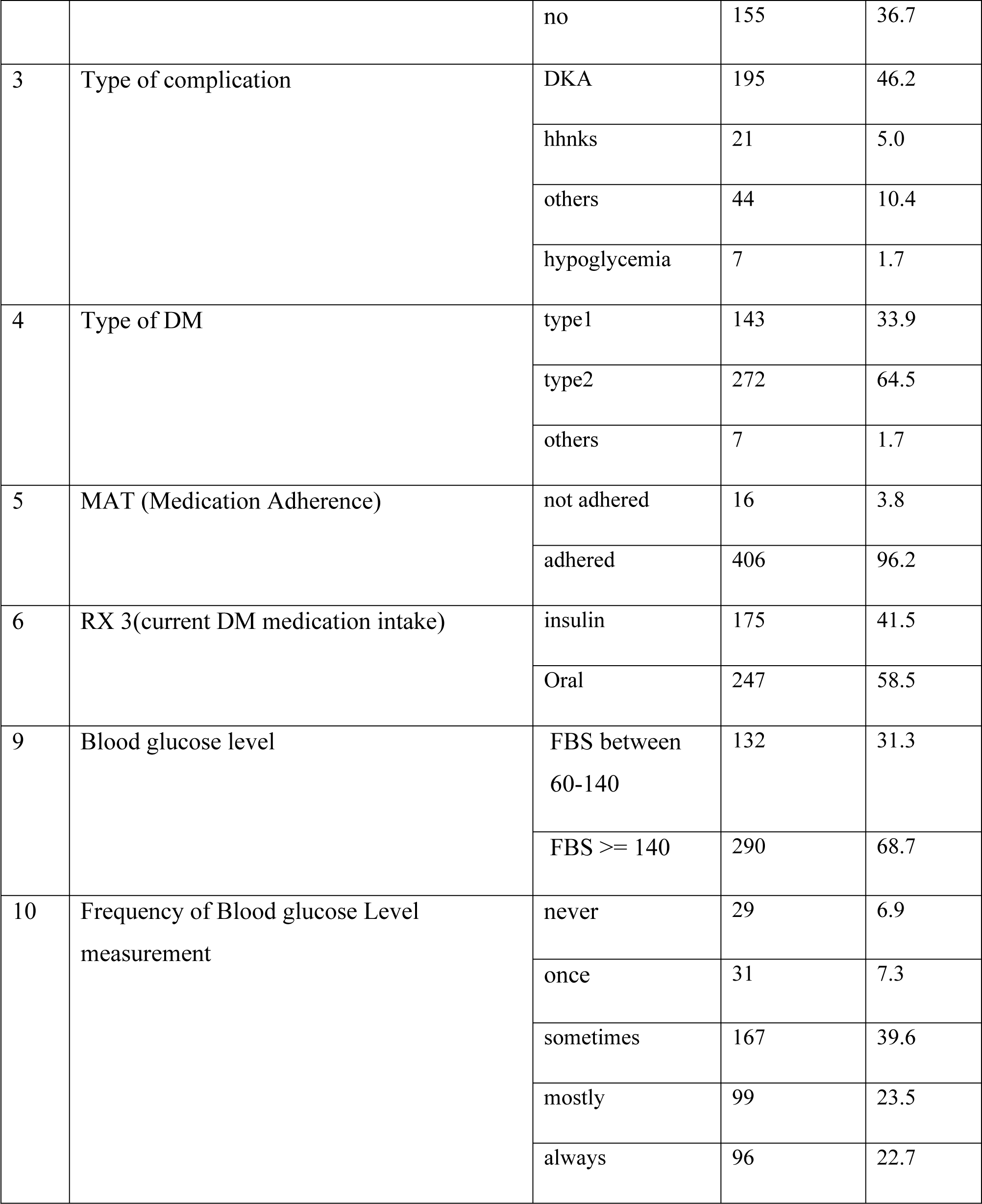
Disease and medication related characteristics for the respondents at selected hospitals of southern Ethiopia, 2021 (n=422).

### Prevalence of treatment outcome

From the total sample the treatment outcome 277 (65.6%) were good. Whereas 145 (34.4%) were found to be poor.

### Sociodemographic factors associated with treatment outcome

From the sociodemographic variables affecting the respondents; only marital status and educational status were found to be associated significantly with the treatment outcome. (See table 4 below).

**Table 4:** - The sociodemographic factors associated with treatment outcome among the respondents at selected hospitals of southern Ethiopia, 2021 (n=422).

| S.n | Variables | category | Treatment outcome |  | Exp(B) | 95% C.I. for EXP(B) |  |
| --- | --- | --- | --- | --- | --- | --- | --- |
|  |  |  | Good | poor |  | Lower | Upper |
| 1 | Age | young | 88(31.8) | 52(35.9) |  |  |  |
|  |  | middle | 90(32.5) | 49(33.8) | 0.921 | 0.565 | 1.502 |
|  |  | old | 99(35.7) | 44(30.3) | 0.752 | 0.459 | 1.232 |
| 2 | Sex | male | 172(62.1) | 92(63.4) | 1.060 | 0.699 | 1.607 |
|  |  | Female | 105(37.9) | 53(36.6) |  |  |  |
| 3 | address | Urban | 190(68.6) | 87(60.0) | 0.687 | 0.452 | 1.043 |
|  |  | Rural | 87(31.4) | 58(40.0) |  |  |  |
| 4 | Marital status | single | 58(20.9) | 24(16.6) | 3.103 | .659 | 14.625 |
|  |  | married | 188(67.9) | 116(80.0) | 4.628 | 1.039 | 20.604 |
|  |  | Widow/divorced | 31(11.2) | 5(3.5) | 1.406 | .206 | 9.619 |
| 5 | Educational status | illiterate | 22(7.9) | 18(12.4) | 1.977 | .949 | 4.120 |
|  |  | Read and write | 39(14.1) | 10(6.9) | .620 | .280 | 1.373 |
|  |  | grade1-8 | 66(23.8) | 47(32.4) | 1.721 | 1.004 | 2.951 |
|  |  | grade10-12 | 63(22.7) | 34(23.4) | 1.304 | .738 | 2.306 |
|  |  | College and above | 87(31.4) | 36(24.8) |  |  |  |
|  |  | low | 69(28.2) | 31(22.0) |  |  |  |
| 7 | Income | middle | 87(35.5) | 58(41.1) | 0.769 | 0.446 | 1.326 |
|  |  | high | 89(36.3) | 52(36.9) | 1.141 | 0.708 | 1.838 |

### Behavioural factors affecting treatment outcome

From the four behavioural variables included in the study; none of them were found to be associated significantly (See table 5 below).

**Table 5:**
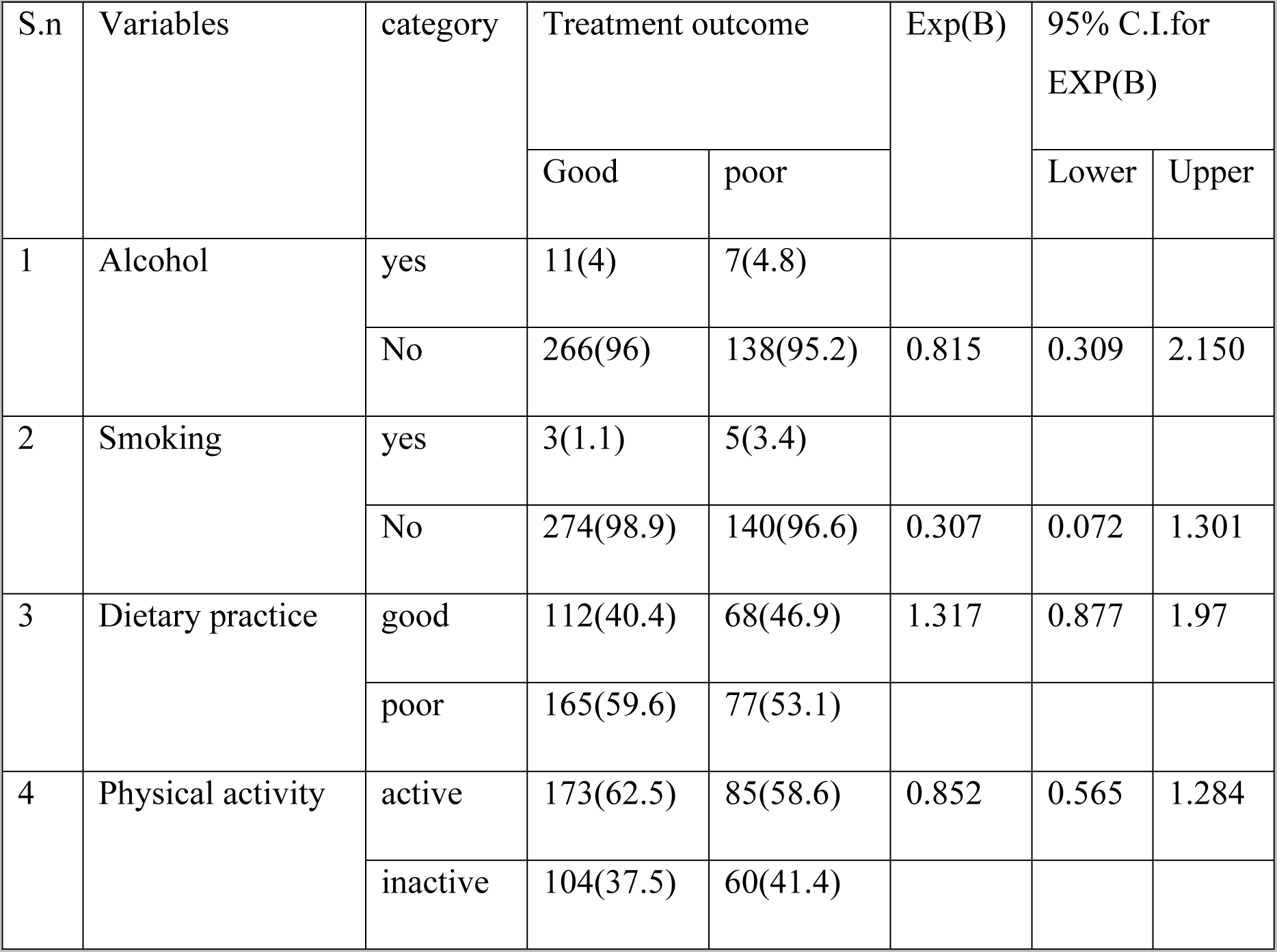
- Behavioural factors associated with treatment outcome among the respondents at selected hospitals of southern Ethiopia, 2021 (n=422).

### Disease and medication related factors associated with treatment outcome

From eight variables studied under disease and medication related factor; the following four factors called Past history of admission, Type of complication, Blood glucose Check-up and Blood glucose level were found to be associated significantly (See table 6 below).

**Table 6:** -behavioural factors associated with treatment outcome among the respondents at selected hospitals of southern Ethiopia, 2021 (n=422).

| S.n | Variables | category | Treatment outcome |  | Exp(B) | 95% C.I.for EXP(B) |  |
| --- | --- | --- | --- | --- | --- | --- | --- |
|  |  |  | Good | poor |  | Lower | Upper |
| 1 | Past history of admission | yes | 164(59.2) | 103(71.0) | 1.69 | 1.098 | 2.601 |
|  |  | No | 100 (41.8) | 55 (29) |  |  |  |
| 2 | Duration of DM | < 10 years | 91 (56.1) | 71 (43.9) | 0.27 | 0.13 | 1.32 |
|  |  | Greater or equal to 10 years | 214(82.3) | 46(17.7) |  |  |  |
| 3 | Presence of complication | yes | 176(63.5) | 91(62.8) | 0.967 | 0.638 | 1.466 |
|  |  | No | 94(36.5) | 61(37.2) |  |  |  |
| 4 | Type of complication | DKA | 121(77.1) | 64(64.6) | 0.465 | 0.259 | 0.833 |
|  |  | hyperglycemia | 36 (22.9) | 35(35.4) |  |  |  |
| 5 | Type of DM | Type I | 90(32.5) | 53(36.6) | 1.472 | 0.276 | 7.857 |
|  |  | Type II | 182(65.7) | 90(62.1) | 1.236 | 0.235 | 6.496 |
|  |  | others | 5(1.8) | 2(1.4) |  |  |  |
| 6 | Medication adherence | yes | 264(95.3) | 142(97.9) |  |  |  |
|  |  | no | 13(4.7) | 3(2.1) | 2.331 | 0.653 | 8.315 |
| 7 | Medication intake | Insulin | 115(41.5) | 60(41.4) |  |  |  |
|  |  | oral | 162(58.5) | 85(58.6) | 1.006 | 0.669 | 1.512 |
| 8 | Blood glucose Check- up/day | yes | 41(68.3) | 19 (31.6) | 0.26 | 0.147 | 0.963 |
|  |  | no | 322 (88.9) | 40(11.1) |  |  |  |
| 9 | Blood glucose level | FBS Between 60-140mg/dl | 118(42.6) | 14(9.7) |  |  |  |
|  |  | FBS >= 140mg/dl | 159(57.4) | 131(90.3) | 0.144 | 0.079 | 0.262 |

From the total of 19 variables studied under this study; only dietary practice, presence of complication and having FBS >= 140 were found to be independently associated with treatment outcome (See table 7).

**Table 7.** Independent predictors predicting treatment outcome among the respondents at selected hospitals of southern Ethiopia, 2021 (n=422).

| Variables | Category | COR :95% C.I. for |  |  | AOR 95% C.I. for |  |  |
| --- | --- | --- | --- | --- | --- | --- | --- |
|  |  | EXP(B) | Lower | Upper | EXP(B) | Lower | Upper |
| Dietary practice | Good | 1.317 | 0.877 | 1.97 | 0.924 | 0.869 | 0.983 |
|  | Poor |  |  |  |  |  |  |
| Presence of complication | yes |  |  |  |  |  |  |
|  | No | 0.967 | 0.638 | 1.466 | 0.425 | 0.217 | .832 |
| FBS | 60-140mg/dl |  |  |  |  |  |  |
|  | >= 140 mg/dl | 0.144 | 0.079 | 0.262 | 0.080 | 0.034 | 0.188 |

## Discussion

According to this study, out of a total sample size of 422 people, 277 (65.6%) had a good treatment outcome, while 145 (34.4%) had a bad treatment outcome. However, it is lower than other studies done in debretabor in Ethiopia (71.4%) (9); Dessie Referral Hospital (70.8%) (10); and Jimma University Teaching Hospital (70.9%) (11). This could be attributed to differences in measuring scales, improved patient treatment outcomes, and research conducted in various health settings in southern Ethiopia.

This study also revealed that those who did not adhere to the suggested dietary practices had a 7.6% higher AOR; 0.924; 95% (0.869, 0.983) risk of receiving a poor treatment outcome than those who did. As a result, this study revealed that dietary recommendations were closely linked to glycemic control in diabetes patients. The findings of this study are consistent with those of studies conducted in Gulf Cooperation Council countries (12) and Turkey (13), which found that low dietary compliance is significantly related to poor glycemic control, and it was also discovered to be associated independently (9).

It is clear that lifestyle changes, such as following the recommended diet, are critical methods for regulating patients’ blood sugar levels. As a result, poor adherence to a recommended diet may make blood sugar control more difficult.

This study also found that individuals who experienced problems had a 57.5% higher AOR; 0.425; and 95% (0.217, 0.832) worse treatment outcomes than those who did not.

Other investigations conducted in Ethiopia’s south west tertiary hospitals (15) and Gondar hospital (14) discovered that complications were significantly associated with poor glycemic outcomes.

Other studies (15) at numerous public hospitals in western Ethiopia found a strong link between the prevalence of complications and poor glycemic control.

When compared to those with FBS between 60 and 140mg/dl, those with FBS >= 140mg/dl had 8% higher AOR; 0.080; and 95% (0.034,0.188) worse treatment outcomes. It is commonly understood that as an individual’s FBS rises, so is the likelihood of a poor treatment outcome.

## CONCLUSION

The magnitude of treatment outcome was determined to be poor in this study. The study also discovered that dietary practices, the existence of complications, and having FBS greater than 140mg/dl all predicted independently.

## RECOMMENDATION

Since the magnitude of treatment outcome were found to be low, the health care providers together with the responsible bodies should strengthen to provide toward improvement of factors affecting the patients by inhibiting them not to be in a state of good treatment.

## LIMITATION S

There could have been recall bias in the prior 7 days while the individuals were answering questions about medication adherence as limitation for this study.

## STRENGTH AND WEAKNESS

The strength of this study was using the standard measurement to measure glycemic control to classify it as poor or good. On the other hand, the weakness of this study is due to its cross-sectional effect of the design

## Data Availability

The data will be deposited in public repositories.

## ACRONYMS AND ABBREVIATION

CBE: Community based education
DM: Diabetes mellitus
HUCSH: Hawassa University Comprehensive Specialized Hospital
WHO: World health organization

## DECLARATION

### Ethics approval and consent to participate

The study was implemented after the ethical review committee of Hawassa University College of Medicine and Health Science approved it. After promising secrecy, each study subject provided written informed consent in order for the study to begin. An individual who was uninterested in participating in the study was permitted to withdraw. We also guarantee that there were no dangers that put the participants in danger.

### Consent for publication

We all agree on publication. I am the corresponding author and the first author of this finding.

### Consent to publish

The consent form is held by the authors. Written informed consent was obtained from the participant.

### Availability of data and materials

The study finding was putted in public repositories. So that the readers can access the data with the following cite called https://www.hindawi.com/research.data/#statement.

### Author’s contribution

The author contributes to the conceptualization and design, data collecting, data analysis and interpretation. They also help to draft articles, revise them, and approve manuscripts before they are published, and they are held accountable for the job they do on the text.

## Acknowledgement

We would like to thank the Hawassa University College of Medicine and Health Science’s Ethical Review Board for conducting an ethical clearance. We would also want to thank the hospitals for agreeing to direct the study in the chronic outpatient clinic. Furthermore, we would like to thank the study participants for their excellent contributions to the study.

Furthermore, the pre-print of this work is available

---

**Figure 1:**
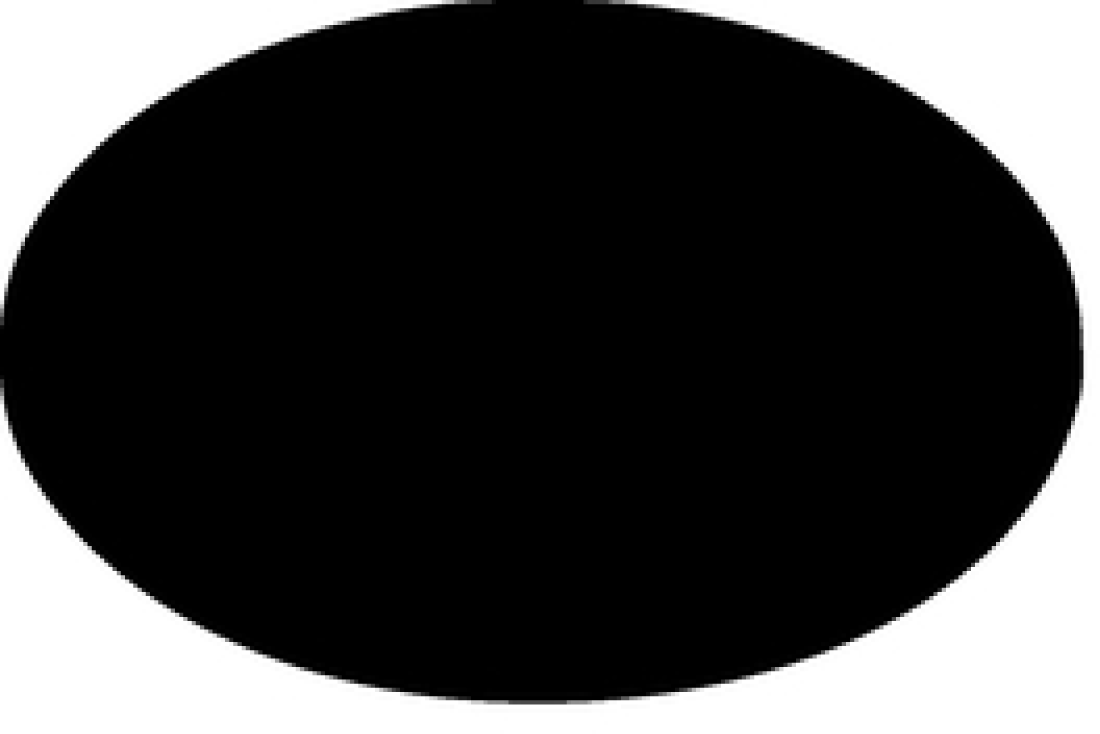
Magnitude of good treatment outcome vs poor treatment outcome Figure

